# Development and Validation of a 12-Item Form of the Participation Questionnaire for Preschoolers

**DOI:** 10.64898/2026.09.10.26362735

**Authors:** Takuto Nakamura, Hirofumi Nagayama

## Abstract

**Aims:** To develop and validate the 12-item short form of the Participation Questionnaire for Preschoolers (PQP-12).

**Methods:** In a cross-sectional study (*n* = 164 aged 3–6 years), 12 items (three per factor) were selected based on item–total correlations while retaining the four-factor structure; confirmatory factor, internal consistency, and construct validity analyses were conducted. In a cohort study (*n* = 275 aged 5–6 years), confirmatory factor and internal consistency analyses were reperformed in an independent sample, and test–retest reliability (intraclass correlation coefficients [ICCs]), measurement error (standard error of measurement [SEM], minimal detectable change [MDC]), and responsiveness were examined.

**Results:** The four-factor model fit well in both samples (comparative fit index = 0.999/0.996) and was superior to the one-factor model. Cronbach’s α was 0.75 and 0.80 (subscales: 0.46– 0.92), with Factor 2’s low value likely reflecting ceiling effects in the older sample. The PQP-12 correlated strongly with the original PQP (*r* = 0.86–0.95); 78% of construct-validity hypotheses and 80% of responsiveness hypotheses were supported. Test–retest reliability was good (ICC = 0.79–0.90), with SEM and MDC being reported.

**Conclusion:** The PQP-12 preserves the original PQP’s structure and measurement properties while reducing respondent burden.

---

Participation of children with autism spectrum disorder (ASD) in daily activities and social situations tends to be restricted owing to difficulty in social communication, restricted interests, repetitive behaviors, and sensory processing features (American Psychiatric Association, 2022; Askari et al., 2015; Nakamura, Dobashi, et al., 2025). Restricted participation among children with ASD has been reported as fewer leisure activities and a bias toward home-based activities, activities with adults, and solitary activities (Askari et al., 2015). Moreover, Nakamura, Nagayama, et al. (2025) showed that participation patterns across participation domains among children with ASD differed from those observed among children with developmental risk without an ASD diagnosis. Thus, participation assessment in children with ASD requires capturing not only reduced participation frequency but also participation patterns characteristic of ASD (Yee et al., 2017).

Participation assessment tools for children with ASD, such as the Structured Preschool Participation Observation adapted for children with ASD based on expert observation (Golos et al., 2022) and the Chinese version of Picture My Participation based on a semi-structured interview (Li et al., 2023), have also been developed; however, these tools require expert involvement or interviews, constraining their utility in practice settings and large-scale surveys. In contrast, the Participation Questionnaire for Preschoolers (PQP) can assess participation without requiring expert involvement. The PQP is a caregiver-report measure consisting of 29 items across four factors, whose internal consistency, construct validity, test–retest reliability, responsiveness, and structural validity have been verified (Nakamura et al., 2024, 2025, 2026). Beyond cross-sectional research, the application of PQP to longitudinal and intervention research is anticipated.

The 29-item length of the PQP does not necessarily impose a small response burden in large-scale surveys. Indeed, quality-of-life and mental-health scales have reduced respondent burden in large-scale longitudinal studies through the development of short forms comprising approximately 10–15 items (e.g., the PedsQL-15) (Chen et al., 2007). Therefore, creating a short form of the PQP with a reduced number of items while retaining its measurement properties is crucial for improving its feasibility in large-scale surveys and longitudinal research.

The present study aimed to develop a 12-item short form of the PQP and to evaluate its reliability and validity

## Methods

The present study used data derived from two observational studies that sought to develop the PQP and evaluate its measurement properties—namely, a cross-sectional study (Study 1) conducted by Nakamura, Nagayama, et al. (2025) and a cohort study (Study 2) by Nakamura et al. (2026). The data of Study 1 were used to assess item selection, structural validity, internal consistency, and construct validity of the short-form PQP, whereas the data of Study 2 were used to re-examine structural validity and internal consistency in an independent sample and to evaluate test–retest reliability and responsiveness. The methods are described separately for each study below.

### Study 1: Cross-Sectional Study

#### Study Design

The cross-sectional study was conducted from December 2021 to February 2023 in Japan and included caregivers of children diagnosed with ASD and children at risk for neurodevelopmental disorders (Nakamura, Nagayama, et al., 2025). In this study, data of 164 children aged 3–6 years who were diagnosed with ASD and had no missing questionnaire data were analyzed. Participant characteristics are summarized in Table 1. Data-collection procedures are detailed in the original study (Nakamura, Nagayama, et al., 2025). This study was conducted in accordance with the Declaration of Helsinki and approved by the Kanagawa University of Human Services Research Ethics Committee (Approval Number: 31-14-009). Informed consent was obtained from all participating caregivers.

**Table 1.** Participant characteristics.

|  | Study 1 | Study 2 |
| --- | --- | --- |
|  | (*n* = 164) | (*n* = 275) |
| Respondent |  |  |
| Relationship (mother) | 153 (93%) | 264 (96%) |
| University graduate | 91 (56%) | 172 (63%) |
| Annual household income |  |  |
| >¥3M | 15 (9%) | 22 (8%) |
| Single-parent household | 8 (5%) | 13 (5%) |
| Target child: demographics |  |  |
| Sex (male) | 124 (76%) | 216 (79%) |
| Age in months | 67.5 (56,74) | 66 (60, 71) |
| Target child: clinical/developmental characteristics |  |  |
| Combined diagnosis |  |  |
| ADHD | 31 (19%) | 57 (21%) |
| ID | 30 (18%) | 74 (27%) |
| Others | 4 (2%) | 13 (5%) |
| SCQ-J score | 10 [6, 15] | – |
| SRS-2-J T-score | – | 78 [69, 87] |

Values are presented as *n* (%) for categorical variables and median [IQR] for continuous variables.
ADHD, attention-deficit/hyperactivity disorder; ID, intellectual disability; SCQ-J, Social Communication Questionnaire – Japanese version; SRS-2-J, Social Responsiveness Scale, Second Edition – Japanese version; ¥, Japanese yen.

### Measures

#### PQP

The PQP is a caregiver-report questionnaire that assesses participation among preschool children with ASD. The original PQP comprises 29 items across four factors (namely, “Friendship and Education,” “Family Satisfaction,” “Daily Living and Independence,” and “Leisure and Community Life”). Each item is rated on a 5-point scale ranging from 1 (“Does not apply”) to 5 (“Applies”), with higher scores indicating better participation. The structural validity, internal consistency (total-score α = 0.83), construct validity, test–retest reliability (total-score intraclass correlation coefficient [ICC] = 0.93), and responsiveness of the PQP have been verified in previous studies (Nakamura et al., 2026; Nakamura, Nagayama, et al., 2025). Because this study was conducted in parallel with the PQP development study, a 36-item prototype was used, from which seven items were subsequently removed (Nakamura et al., 2024). The removed items were excluded from the final scoring.

### Social Communication Questionnaire – Japanese Version (SCQ-J)

The SCQ-J is a caregiver-completed questionnaire designed to evaluate social and communication features, which are considered core ASD symptoms. The SCQ-J can be used for individuals aged ≥4 years. In this study, the Lifetime Form of the SCQ-J was used. Its internal consistency (α = 0.926) and test–retest reliability (ICC = 0.961) have been verified (Uchiyama, 2015a, 2015b). Higher scores indicate stronger ASD symptoms.

### Short Sensory Profile (SSP)

The SSP is a caregiver-report questionnaire assessing sensory features. In this study, the Japanese version of the SSP was used (Dunn, 1999; Tanii et al., 2015). Its internal consistency (Cronbach’s α = 0.537–0.880) and construct validity (verified through correlations with intelligence quotient, autism symptoms, and adaptive behaviors) have been confirmed (Tanii et al., 2015). Higher scores indicate stronger atypical sensory features in children.

### Survey of Family Environment – Japanese Version (SFE-J)

The SFE-J is a 30-item questionnaire that measures family functioning, including the environment inside and outside the family. In this study, the total score indicating overall satisfaction with family functioning was used, following the method employed in the original PQP development study (Nakamura, Nagayama, et al., 2025). Its internal consistency (Cronbach’s α = 0.94), test–retest reliability (ICC = 0.92), and structural validity (confirmatory factor analysis) have been verified (Hohashi & Honda, 2012). Higher scores indicate better family functioning.

### Age

Data on children’s age (in months) were collected from caregiver reports.

### Data Analysis

#### Development of the Short-Form PQP

First, the number of items for the short-form PQP was considered. Given that participation is a multidimensional context-dependent construct, excessive item reduction may compromise the scale’s content validity, particularly the comprehensiveness required to sufficiently cover the construct to be measured (Terwee et al., 2018). Therefore, the four-factor structure of the PQP was retained (Nakamura, Nagayama, et al., 2025) to ensure the multidimensionality of participation.

Next, the top four items with the highest item–total correlations were extracted for each factor. Subsequently, one item was removed for each factor to minimize overlap in content domains while maximizing the coverage of participation domains (Nakamura et al., 2024), to reduce items with ceiling or floor effects (mean ± 1 standard deviation [SD] ≥5 or ≤1), and to avoid bias toward any of the three PQP sections (“the child’s behavior,” “the parent’s feelings,” and “the child’s feelings”). Finally, a 12-item short-form of the PQP (PQP-12) was selected.

### Structural Validity

Confirmatory factor analysis was conducted to examine the structural validity of the PQP-12. Twelve selected items were specified based on the *a priori* four-factor structure, with each item being loaded on only its corresponding single factor. Correlations among factors were freely estimated. Weighted least squares means and variance adjusted (WLSMV) estimation, which is suitable for ordinal data, was performed. Model fit was assessed using the comparative fit index (CFI), Tucker–Lewis index (TLI), root mean square error of approximation (RMSEA), and standardized root mean square residual (SRMR), with CFI and TLI ≥0.90 (≥0.95 indicating good fit), RMSEA ≤0.08 (≤0.06 indicating good fit), and SRMR ≤0.08 being specified as indices of fit. Additionally, a one-factor model was estimated as a competing model to examine structural validity, and its fit was compared. These fit cutoffs were proposed under maximum likelihood estimation assuming normality and continuous data (Hu & Bentler, 1999). WLSMV estimation for ordinal data tends to overestimate CFI and TLI and underestimate RMSEA, making model fit prone to optimistic evaluation (Xia & Yang, 2019). Therefore, in this study, the difference in fit between the four-factor and one-factor models (ΔCFI) was emphasized using ΔCFI >0.01 as an index of a meaningful difference between the two models (Cheung & Rensvold, 2002) instead of relying solely on the absolute values of fit indices.

### Internal Consistency and Construct Validity

Internal consistency of the PQP-12 total and factor scores was evaluated using Cronbach’s α. Based on the previous study (Nakamura, Nagayama, et al., 2025), construct validity was examined by setting *a priori* hypotheses for the correlations between the PQP-12 total score and the following variables: age in months (no to weak positive correlation, *r* = 0.0–0.4), SCQ-J score (weak to moderate negative correlation, |*r*| = 0.2–0.7), SSP score (moderate negative correlation, |*r*| = 0.4–0.7), and SFE-J score (moderate positive correlation, *r* = 0.4–0.7). Furthermore, correlations between the PQP-12 and original PQP total and factor scores were hypothesized to be ≥0.70. All hypotheses were tested using Spearman’s rank correlation coefficient, as in the original PQP development study. Each hypothesis was deemed to be supported if the observed correlation coefficient met the conditions specified in the hypothesis. Construct validity was considered sufficient when at least 75% of all hypotheses were supported (Prinsen et al., 2018).

### Study 2: Cohort Study

The cohort study used data from an existing cohort study evaluating the reliability, responsiveness, and interpretability of the PQP in children with ASD (Nakamura et al., 2026). Participant characteristics are summarized in Table 1. Data-collection procedures are detailed in the original study (Nakamura et al., 2026). The cohort study was approved by the same ethics committee (Approval Number: 31-14-009) and was preregistered before data analysis (UMIN000054162). Informed consent was obtained through the online platform.

### Measures

#### PQP

The PQP used in Study 2 was the same 36-item prototype utilized in Study 1 (Nakamura et al., 2024). In Study 2, the total score and subdomain scores for each factor were computed from the PQP-12 items developed in Study 1 and were used in the analyses.

### Social Responsiveness Scale, Second Edition – Japanese version (SRS-2-J)

The SRS-2-J is a caregiver-report questionnaire designed to measure the severity of ASD symptoms. In this study, participant characteristics were described using the sex-normed T-score of the child version (Constantino & Gruber, 2017). Internal consistency was high in the present data (Cronbach’s α = 0.93). Higher scores indicate stronger ASD symptoms.

### Strengths and Difficulties Questionnaire (SDQ)

The SDQ is a questionnaire that assesses emotional and behavioral difficulties among children. In this study, the Japanese parent-report version for ages 4–16 years was used (Matsuishi et al., 2008). Three indices were employed: the total difficulty score, peer problems, and prosocial behavior. Internal consistency has been reported as acceptable for the total difficulty score (Cronbach’s α = 0.77), low for peer problems (α = 0.52), and somewhat low for prosocial behavior (α = 0.69). Higher scores for all indices except for prosocial behavior indicate greater difficulties; for prosocial behavior, higher scores suggest better adaptive functioning.

### Revised Family Outcomes Survey – Japanese version (FOS-J)

The FOS-J evaluates the impact of support or intervention programs on children and families (Ueda et al., 2015). In this study, only the family-outcome domain, which comprises five subscales, was used. The overall internal consistency of the FOS-J is very high (Cronbach’s α = 0.94) (Ueda et al., 2015). Higher scores indicate that support services generate more beneficial outcomes for the family.

### Global Rating of Change (GRC) Scale

The GRC Scale is an anchor measure on which respondents subjectively rate changes in status associated with intervention or the passage of time (Kamper et al., 2009; Revicki et al., 2008). In this study, test–retest reliability was assessed using only GRC Scale 1, a single item on changes in a child’s life compared with that at one week earlier, which was rated on a 7-point scale ranging from 1 (“Very much worse”) to 7 (“Very much improved”) (Nakamura et al., 2026).

### Statistical Analysis

#### Re-examination of Structural Validity and Internal Consistency

Confirmatory factor analysis was conducted using the same procedure as in Study 1 (four-factor model, each item loaded on its corresponding single factor, factor correlations freely estimated, WLSMV estimation for ordinal data). A one-factor model was compared as a competing model. The fit criteria were the same as in Study 1. Internal consistency was examined by computing Cronbach’s α for the total score and each factor.

### Test–Retest Reliability

Retest was administered at one week after T1 (at T2). ICCs for the PQP-12 total and factor scores were computed for participants who responded “4: no change” on the GRC Scale. ICC ≥0.70 was applied as the criterion for sufficient test–retest reliability (Terwee et al., 2007). Standard error of measurement (SEM) was calculated as follows: SEM = SD × √(1 − ICC). Minimal detectable change (MDC), which represents the smallest change detectable beyond measurement error, was calculated as follows: MDC = 1.96 × SEM × √2.

### Responsiveness

To examine the responsiveness of the PQP-12, based on the *a priori* hypotheses of the previous study (Nakamura et al., 2026), correlations were computed between the change scores from Time 1 to Time 3 and from Time 3 to Time 4—each representing change over approximately three months—and each SDQ and FOS index: SDQ total difficulty score (weak to moderate negative correlation, |*r*| = 0.2–0.5), SDQ peer problem score (weak negative correlation, |*r*| = 0.1–0.3), SDQ prosocial behavior score (weak to moderate positive correlation, *r* = 0.2–0.5), and FOS-J total score (weak positive correlation, *r* = 0.0–0.3). Furthermore, correlations between the total-score change scores of the PQP-12 and the original PQP were hypothesized to be ≥0.70 at each interval. All hypotheses were tested using Spearman’s rank correlation coefficient, as in the original PQP development study. Each hypothesis was deemed to be supported if the observed correlation coefficient met the conditions specified in the hypothesis. Responsiveness was considered sufficient when at least 75% of the hypothesis tests were supported (Prinsen et al., 2018).

### Use of Generative Artificial Intelligence

In preparing this manuscript, the authors used a generative AI tool, Claude (Anthropic; Claude Fable 5), for language editing and for checking the consistency of the reported statistics. All analyses, interpretations, and the final content were verified and approved by the authors, who take full responsibility for the content of the manuscript.

## Results

### Study 1: Cross-Sectional Study

#### Development of the PQP-12

Item–total correlations for each factor are presented in Table 2. For Factor 1, item 6, which reflected broader participation, was retained, whereas item 7, which depended on a specific play format, was removed. For Factor 3, item 28, which reflected a child’s perspective, was retained, whereas item 2, which overlapped in content and showed a ceiling effect, was removed. For Factors 2 and 4, items were selected in order of the highest item–total correlations (Table 2).

**Table 2.**
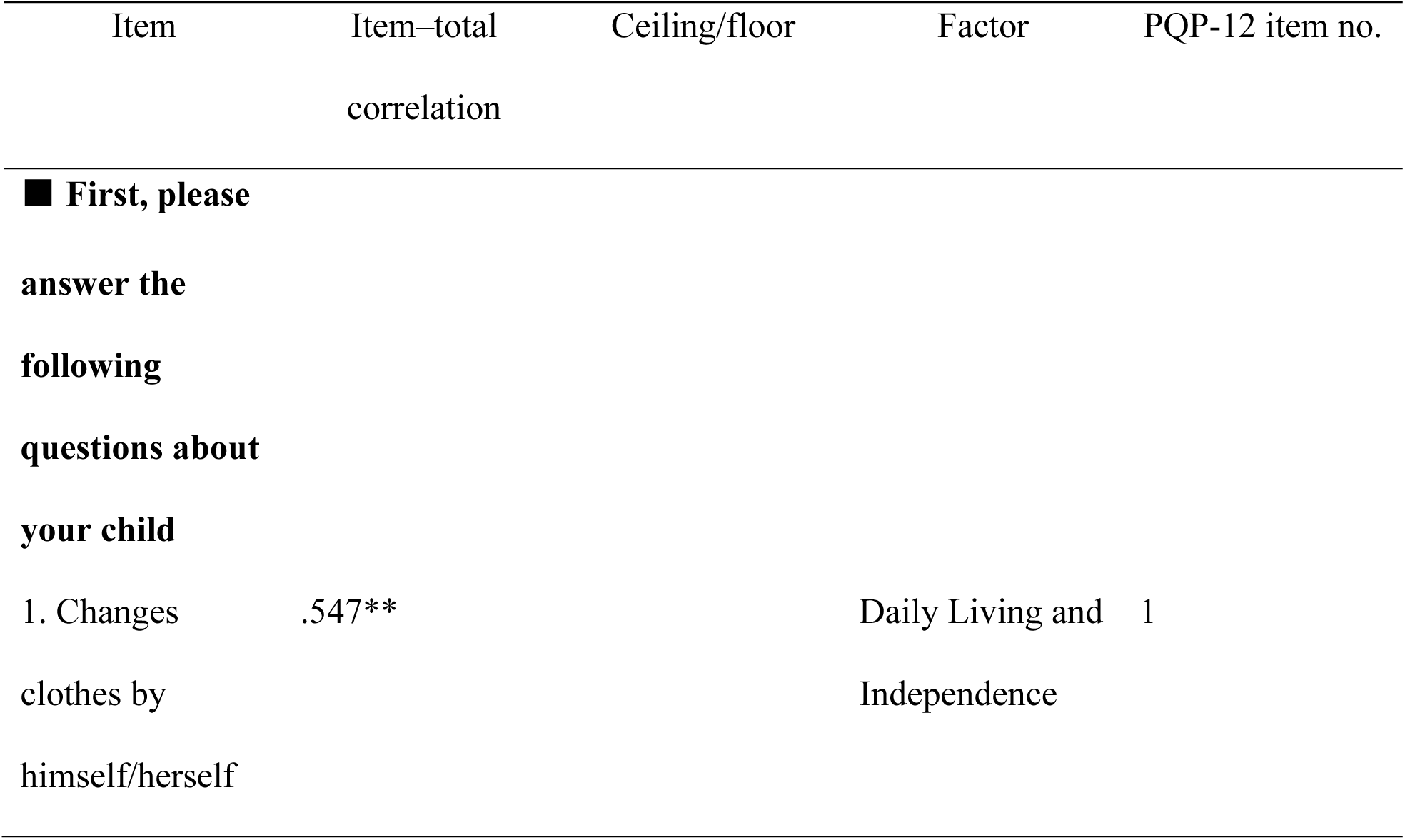
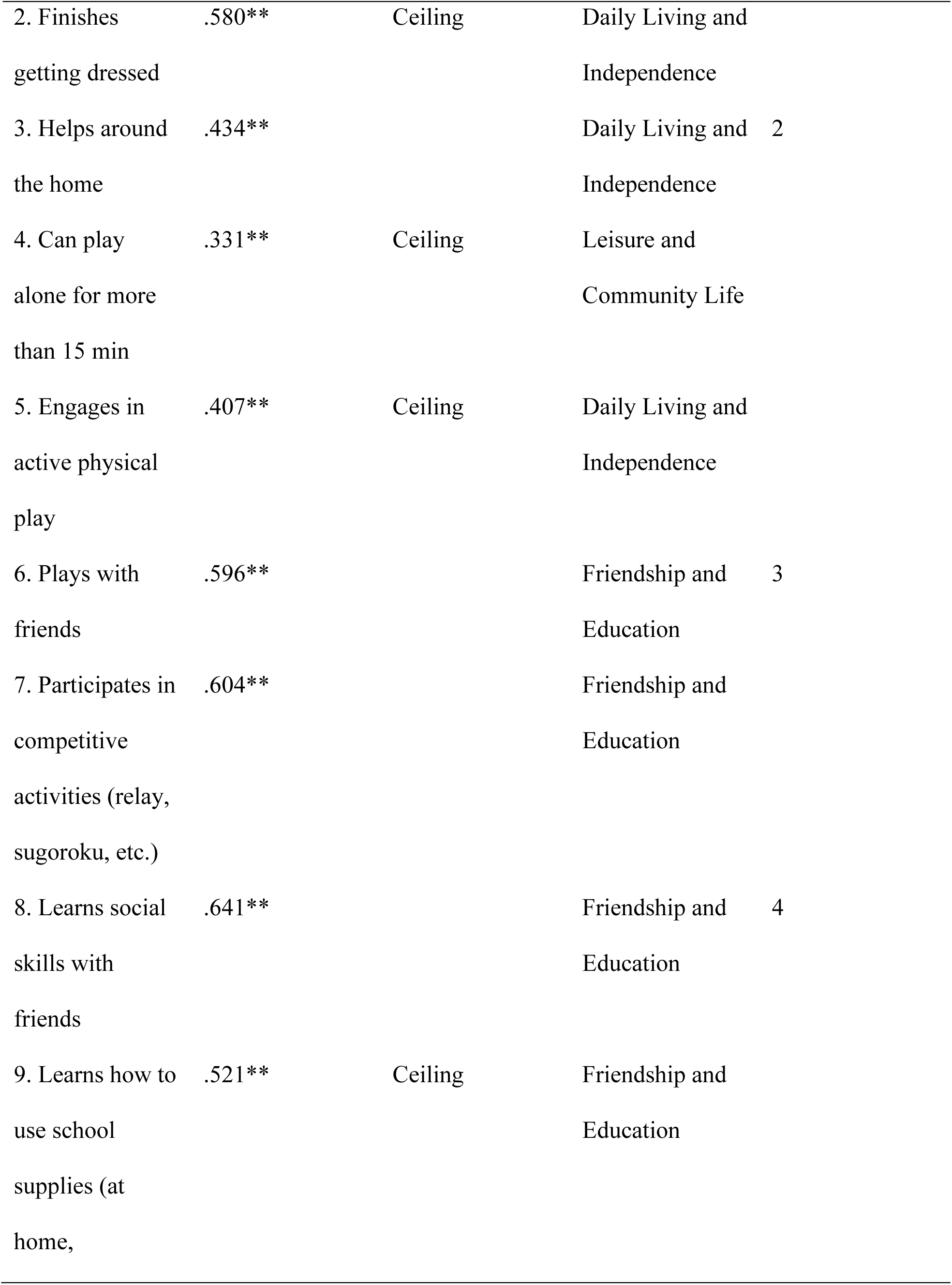
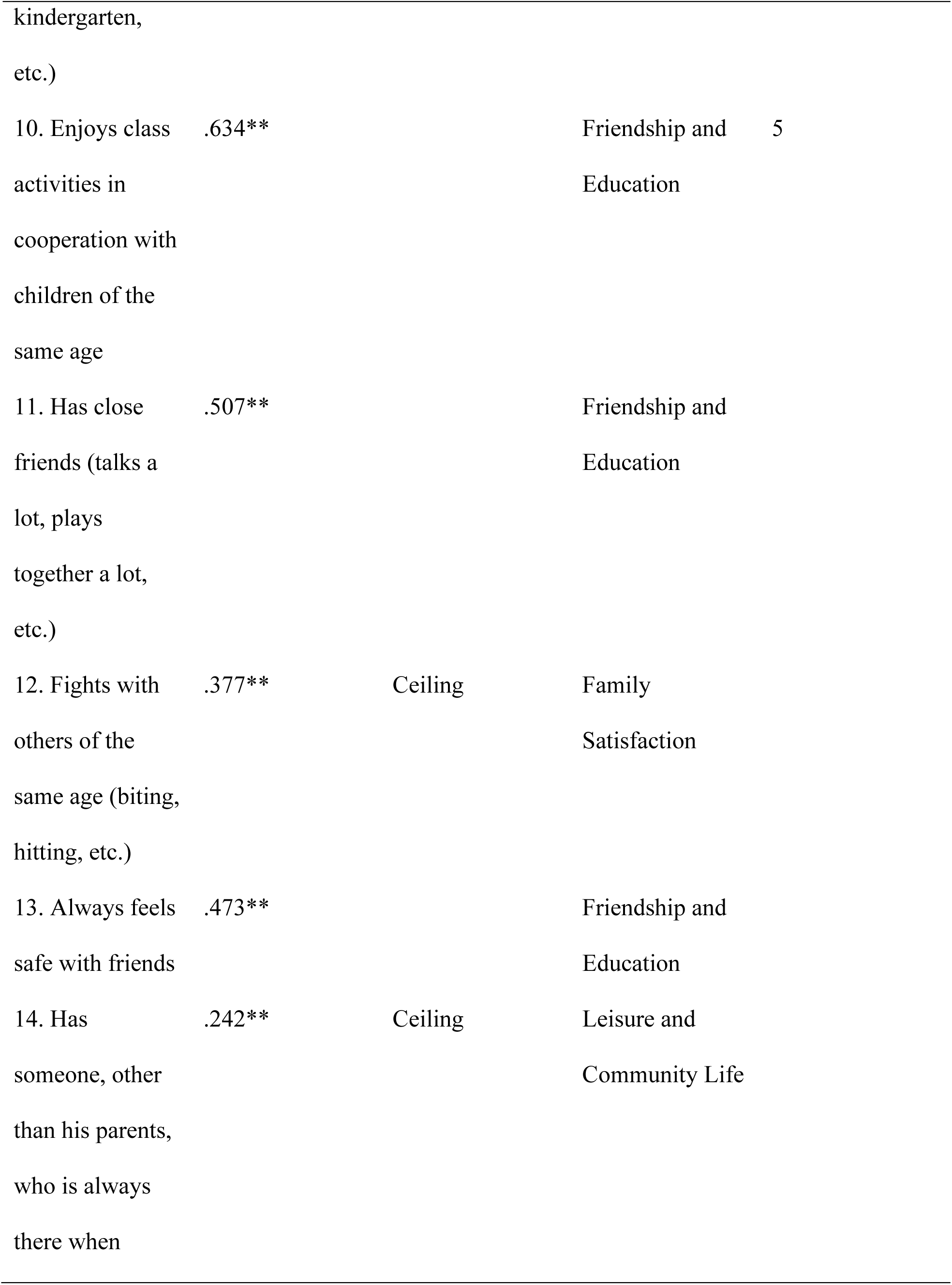
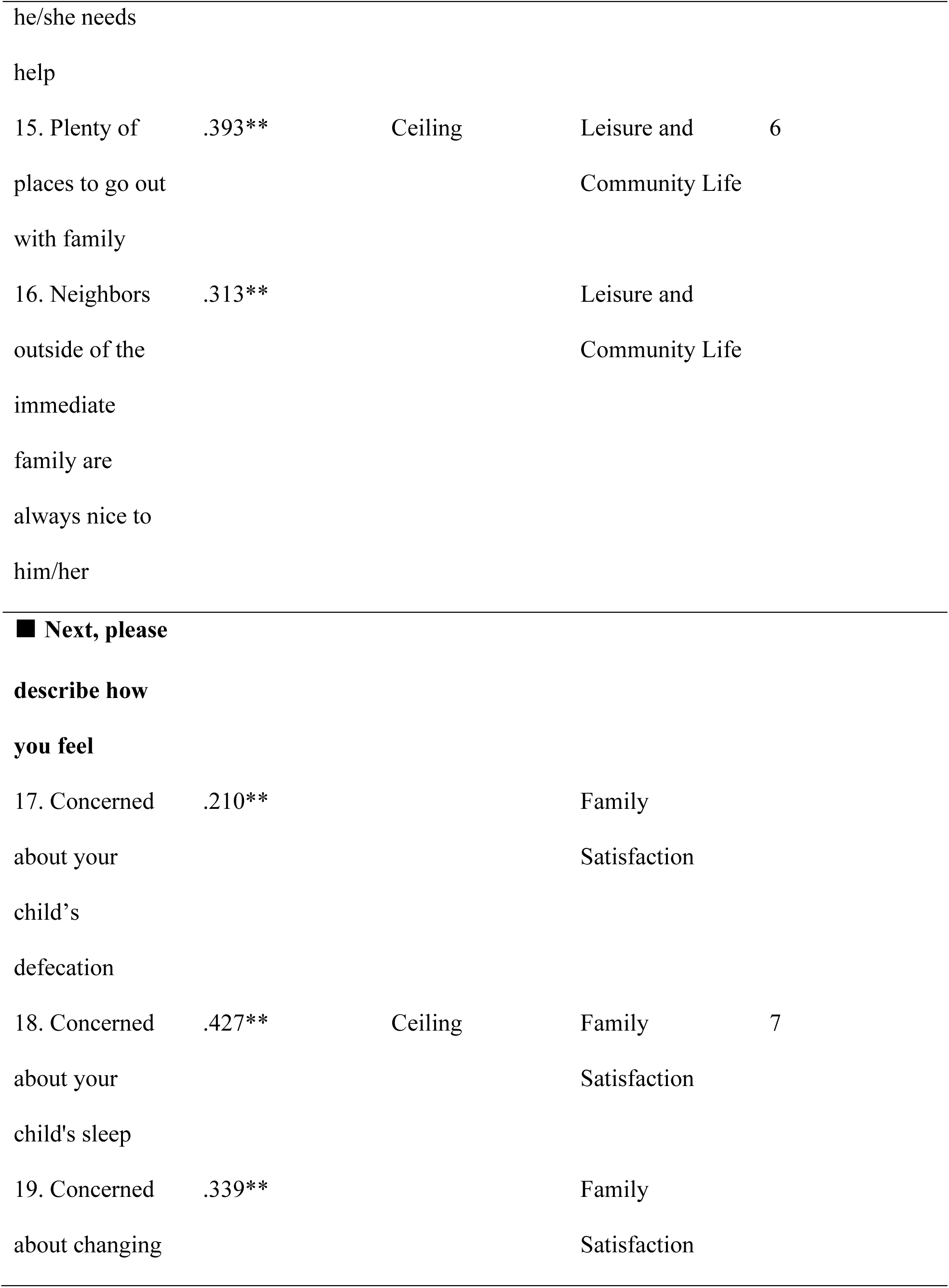
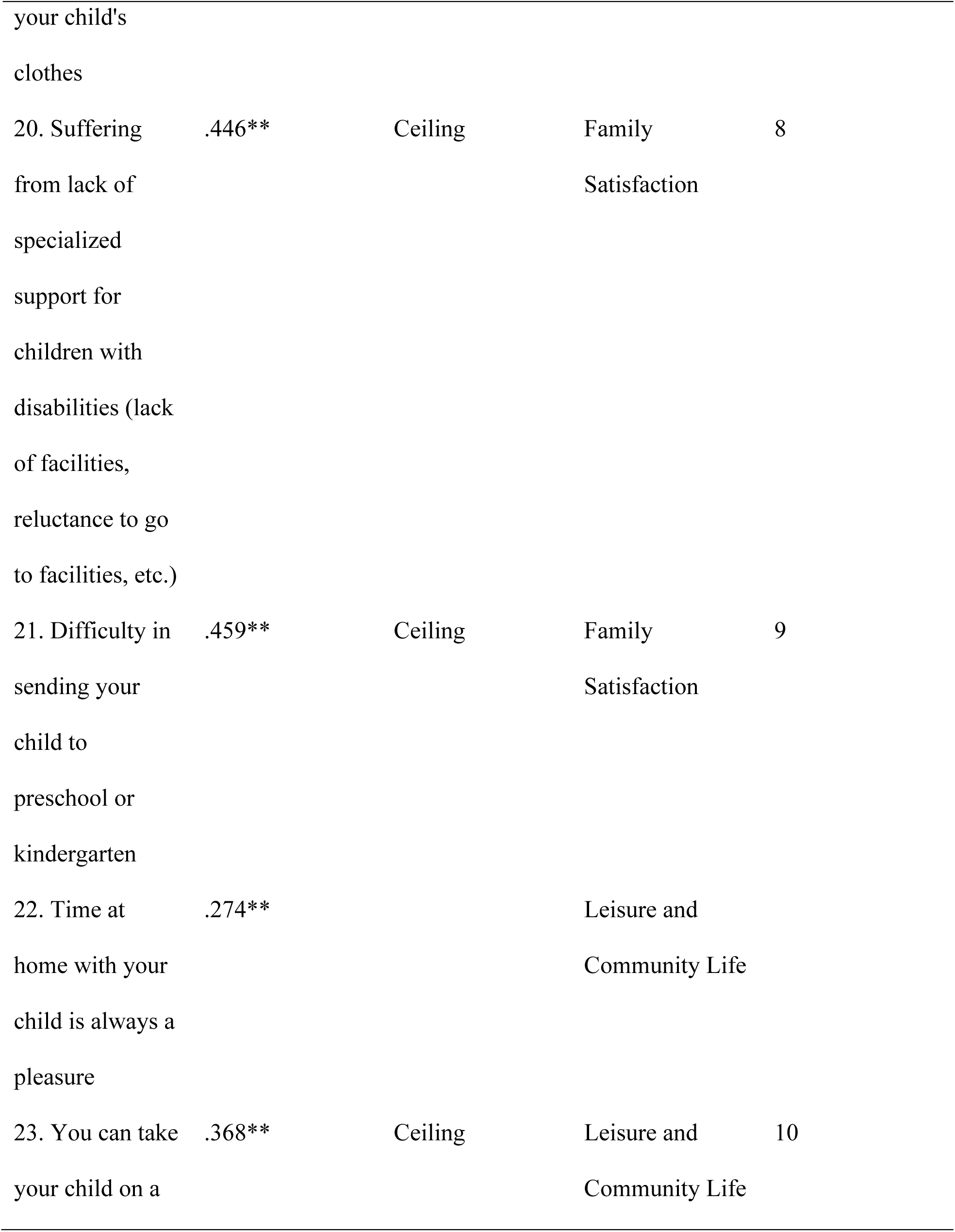
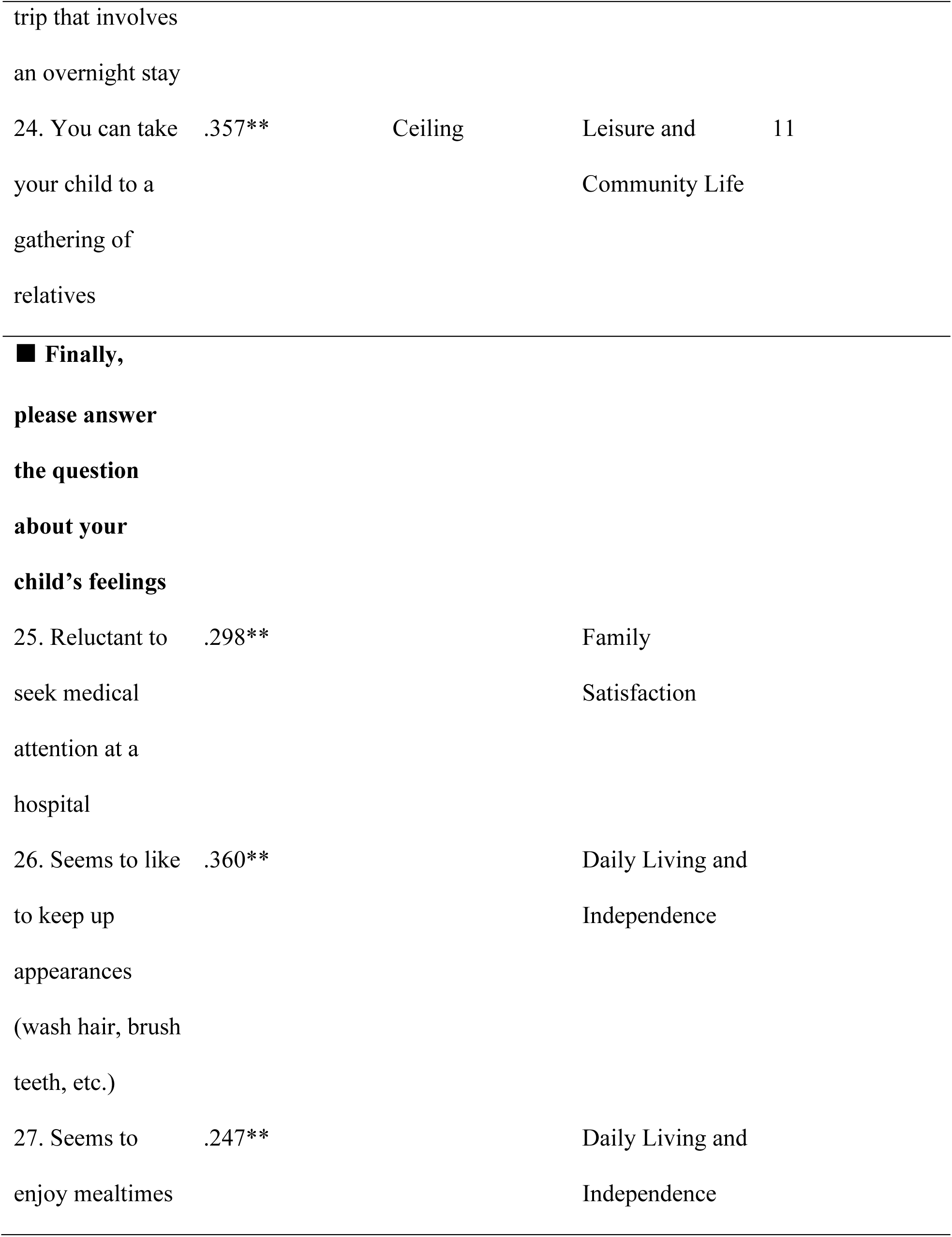
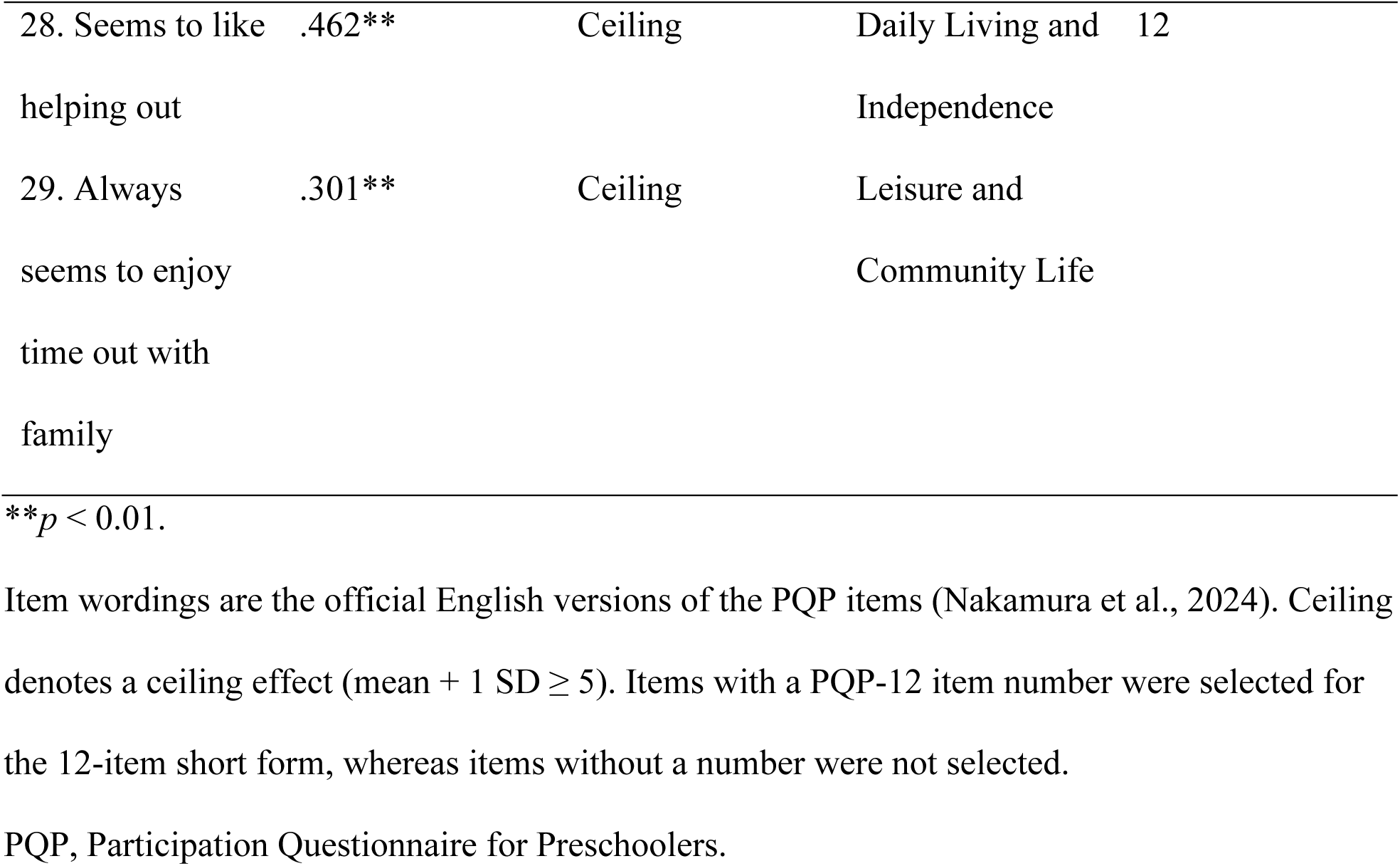
Item–total correlations of the original PQP and item selection for the 12-item short form (PQP-12).

#### Structural Validity

The four-factor model fit well (CFI = 0.999, RMSEA = 0.014), whereas the one-factor model fit poorly (CFI = 0.738, RMSEA = 0.262). Furthermore, the four-factor model was superior to the one-factor model (ΔCFI = 0.262). All standardized factor loadings (range: 0.60–0.89) were significant (*p* < .001). Inter-factor correlations ranged from 0.07 to 0.50 (Table 3).

**Table 3.** Structural validity and internal consistency of the PQP-12: comparison between Study 1 and Study 2.

| | Study 1<br>($n = 164$) | Study 2<br>($n = 275$) | Criterion |
| --- | --- | --- | --- |
| Confirmatory factor analysis (four-factor model) |  |  |  |
| CFI | .999 | .996 | $\geq .95$ good |
| TLI | .999 | .994 | $\geq .95$ good |
| RMSEA | .014 | .048 | $\leq .06$ good |
| SRMR | .069 | .060 | $\leq .08$ |
| Standardized loadings (range) | .600–.886 | .495–.957 | All $p < .001$ |
| Factor correlations<br>(range) | .071–.497 | .340–.577 |  |
| One-factor model<br>(CFI/RMSEA) | .738/.262 | .926/.193 |  |
| ΔCFI (four- vs.<br>one-factor) | .262 | .070 | $\geq .01$ favors four-factor |
| Cronbach's $\alpha$ | | | |
| Total score | .748 | .795 | $\geq .70$ |
| Factor 1 | .855 | .917 |  |
| Factor 2 | .812 | .461 |  |
| Factor 3 | .684 | .682 |  |
| Factor 4 | .715 | .707 |  |

Study 1, cross-sectional sample; Study 2, cohort sample (baseline assessment).
Confirmatory factor analyses were conducted with the WLSMV estimator treating items as ordinal. The four-factor model specified each item on its corresponding factor, with factor correlations freely estimated.
CFI, comparative fit index; TLI, Tucker–Lewis index; RMSEA, root mean square error of approximation; SRMR, standardized root mean square residual; ΔCFI, difference in CFI between the four-factor and one-factor models. The lower $\alpha$ for Factor 2 in Study 2 reflects range restriction (ceiling effects) in the older (5–6-year) cohort; test–retest reliability for Factor 2 remained good (ICC = 0.80).

#### Internal Consistency and Construct Validity

The PQP-12 exhibited generally acceptable internal consistency, with Cronbach’s α of 0.75 for the total score and 0.68–0.86 for the factors. With respect to construct validity, seven (78%) out of nine *a priori* hypotheses were supported (Table 4). Correlations with age and SCQ-J were within hypothesized ranges. In contrast, correlations with the SSP (*r* = −0.38) and SFE-J (*r* = 0.12) did not reach the lower bounds of hypothesized ranges. Additionally, the correlation with the SFE-J was below the hypothesized range for the original PQP (*r* = 0.15). All correlations with the original PQP were ≥0.70 (*r* = 0.86–0.95), with all hypotheses being supported.

**Table 4.** Hypothesis testing results for construct validity.

|  | PQP-12 | Original PQP<br>(Nakamura et al. 2025) |
| --- | --- | --- |
| PQP | 0.926** |  |
| Child's age (in months) | 0.213** | 0.215** |
| SCQ-J | -0.638** | -0.608** |
| SSP | -0.376** | -0.415** |
| SFE-J | 0.117 | 0.149 |

\*\* $p < 0.01$ , \* $p < 0.05$ .
SCQ-J, Social Communication Questionnaire – Japanese version; SSP, Short Sensory Profile; SFE-J, Survey of Family Environment – Japanese version.

### Study 2: Cohort Study

#### Participant Characteristics

Participant characteristics of the analytic sample in Study 2 are summarized in Table 1.

### Re-examination of Structural Validity and Internal Consistency

In the Study 2 sample (*n* = 275), the four-factor model fit well (CFI = 0.996, RMSEA = 0.048), whereas the one-factor model fit poorly (CFI = 0.926, RMSEA = 0.193; ΔCFI = 0.070). All standardized factor loadings (range: 0.50–0.96) were significant (*p* < .001). Inter-factor correlations ranged from 0.34 to 0.58. Cronbach’s α was 0.80 for the total score and 0.46–0.92 for the factors, with Factor 2 being the lowest (Table 3). Three items in Factor 2 had higher means and smaller SDs in Study 2 (range of item means: 3.7–4.0, range of SDs: 1.2–1.5) compared with Study 1 (range of item means: 3.0–3.1, range of SDs: 1.6–1.7), and their inter-item correlations decreased from 0.54–0.67 to 0.19–0.29.

### Test–Retest Reliability, Measurement Error, and Interpretability

ICCs were 0.90 for the total score, 0.90 for Factor 1, 0.80 for Factor 2, 0.79 for Factor 3, and 0.88 for Factor 4 (Table 5). The SEM and MDC for the total score were 2.73 and 7.57 points, respectively.

**Table 5.**
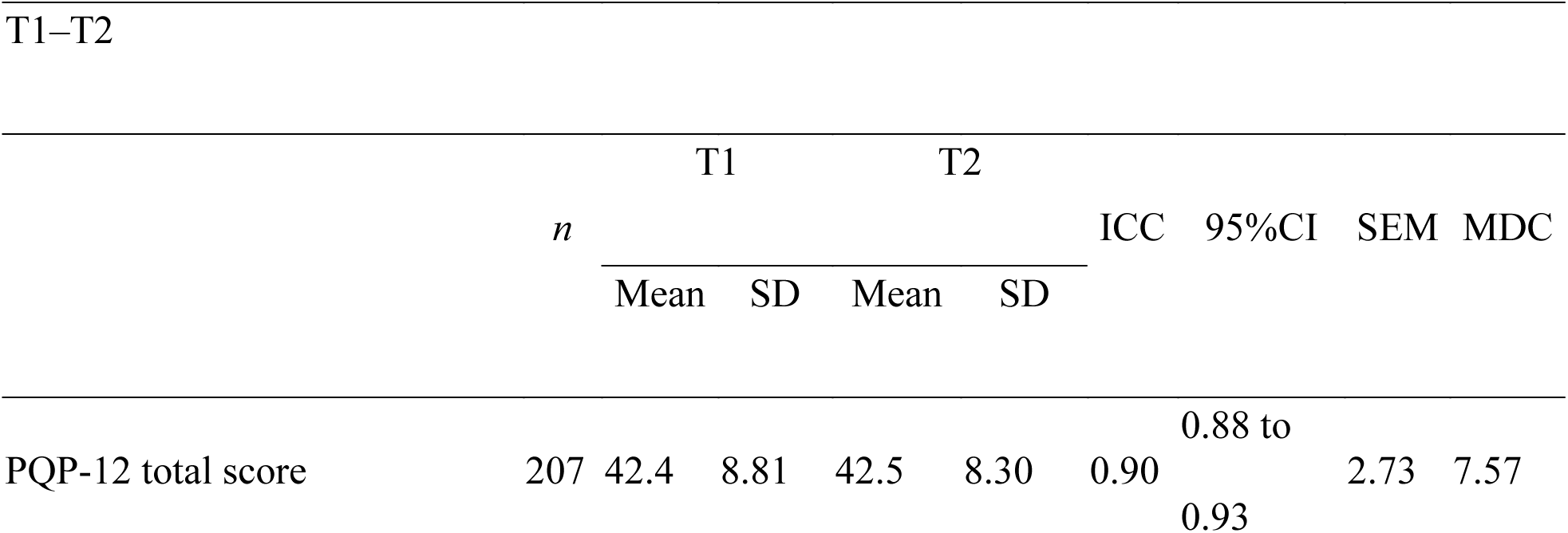
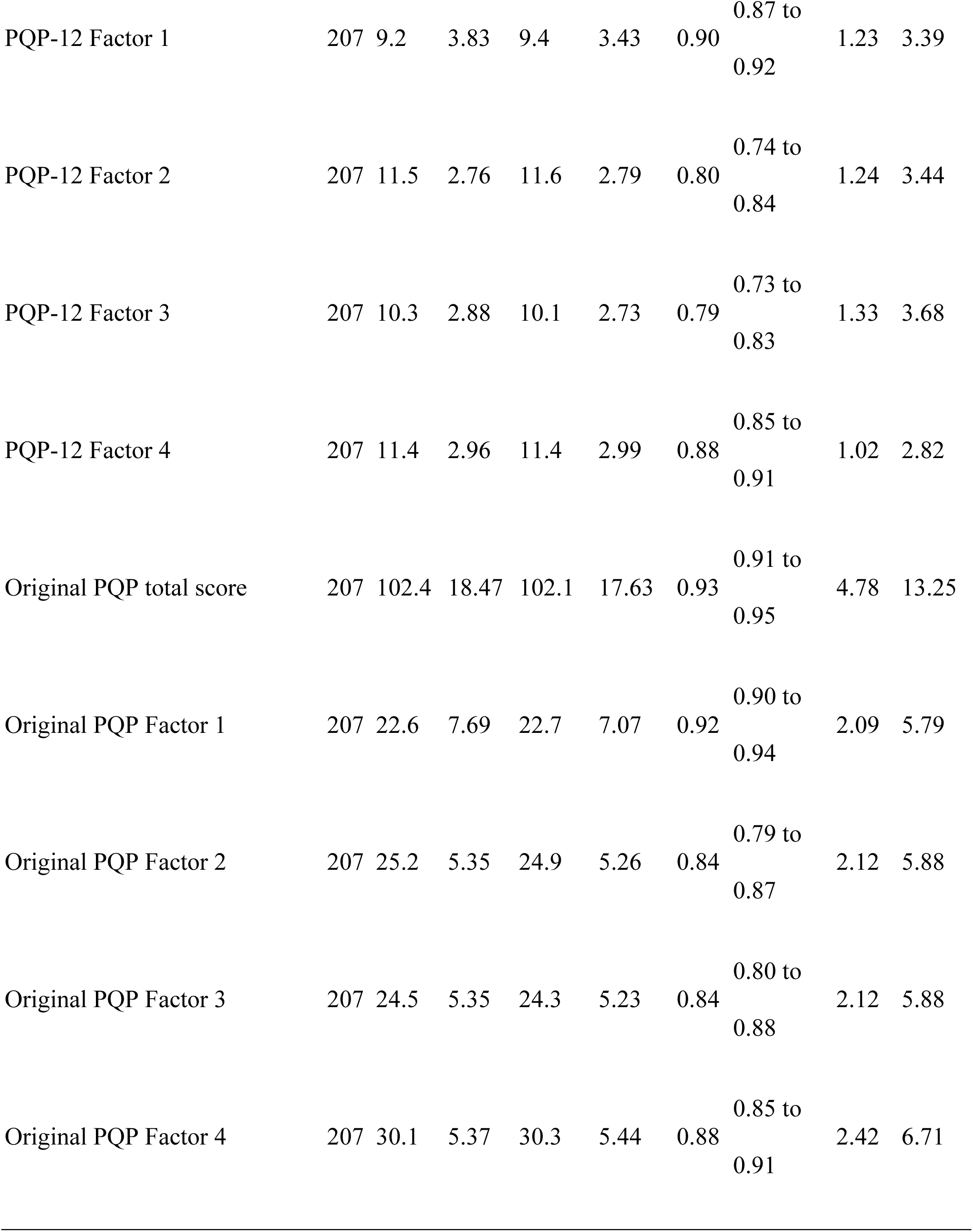
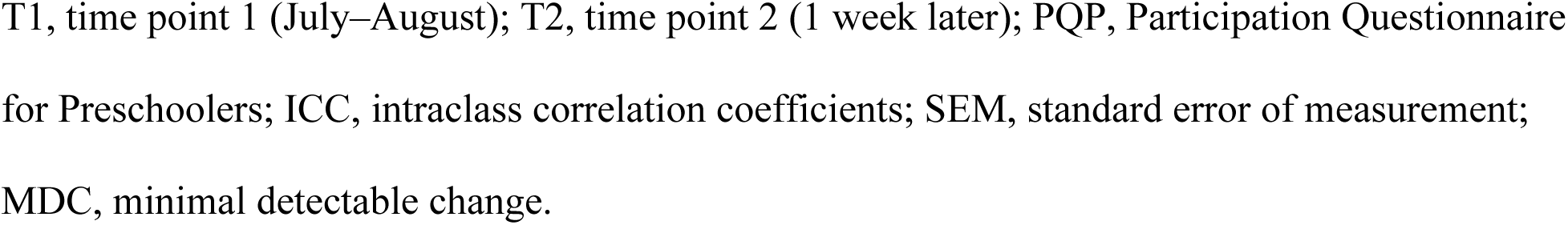
Test–retest reliability and standard error of measurement verification results.

### Responsiveness

Regarding responsiveness, eight (80%) out of ten hypothesis tests were supported (Table 6). Correlations with the SDQ total difficulty score, SDQ peer problem score, and FOS total score were within hypothesized ranges at both intervals. In contrast, correlations with the SDQ prosocial behavior score (0.12/0.19) did not reach the lower bound of the hypothesized range at either interval. Notably, this hypothesis was not supported for the original PQP (0.11/0.18). Correlations between PQP-12 and original PQP total-score changes were 0.83 (T1–T3) and 0.80 (T3–T4), both supporting the hypotheses.

**Table 6.** Hypothesis testing results for responsiveness validation.

| T1–T3 | PQP-12 | Original PQP | Expected correlation (hypothesis) |
| --- | --- | --- | --- |
|  |  | (Nakamura et al. 2026) |  |
| SDQ-TDS | −0.27** | −0.35** | Weak–moderate negative (−0.2 to −0.5) |
| SDQ-PP | −0.30** | −0.28** | Weak negative (−0.1 to −0.3) |
| SDQ-PB | 0.12 | 0.11 | Weak–moderate positive (0.2 to 0.5) |
| FOS-J | 0.13* | 0.05 | Weak positive (0.0 to 0.3) |
| Change-score |  |  | ≥0.70 |
| correlation with the original PQP | 0.83** | — |  |

| T3–T4 | PQP-12 | Original PQP | Expected correlation (hypothesis) |
| --- | --- | --- | --- |
|  |  | (Nakamura et al.<br>2026) |  |
| SDQ-TDS | −0.27** | −0.42** | Weak–moderate negative (−0.2 to −0.5) |
| SDQ-PP | −0.21** | −0.26** | Weak negative (−0.1 to −0.3) |
| SDQ-PB | 0.19** | 0.18** | Weak–moderate positive (0.2 to 0.5) |
| FOS-J | 0.15* | 0.13* | Weak positive (0.0 to 0.3) |
| Change-score |  |  | ≥0.70 |
| correlation with<br>the original PQP | 0.80** | — |  |

\*\* $p < 0.01$ , \* $p < 0.05$ . All correlation coefficients are Spearman's rho. Change-score correlation rows show Spearman correlations between the total-score change scores of the PQP-12 and the original PQP (hypothesis: $\geq 0.70$ ).
PQP, Participation Questionnaire for Preschoolers; SDQ, Strengths and Difficulties Questionnaire; FOS-J, Revised Family Outcomes Survey – Japanese version.

## Discussion

In the present study, we developed the PQP-12, a 12-item short form of the PQP, using data from two observational studies and evaluated its measurement properties. We discuss each measurement property in turn below.

With respect to structural validity, the PQP-12 showed high structural validity as a four-factor scale. The four-factor model fit well and was clearly superior to the one-factor model, indicating that the scale has a multidimensional structure rather than a single construct. This four-factor structure was also well reproduced in the Study 2 sample, which was independent of the sample used for item selection, suggesting that the structural validity of the short form is not attributable to overfitting from item selection.

Regarding internal consistency, the total score and Factors 1, 3, and 4 were at generally acceptable internal consistency levels in both samples and were stably reproduced in the independent sample. In Study 2, the internal consistency coefficient for Factor 2 decreased because the Study 2 sample was biased toward the upper end of the age range (5–6 years), in which the score distribution of Factor 2 items was compressed upward and the inter-item variance was reduced (range restriction associated with a ceiling effect). However, Factor 2 showed good test–retest reliability and was established as an independent factor in the confirmatory factor analysis (i.e., low α reflects the attenuation of inter-item correlations). Because the reproducibility of measurement and the factor structure are preserved, it is interpreted not as an intrinsic defect of the scale but as a limited finding that reflects range restriction in an age-biased sample.

As for construct validity, 78% of *a priori* hypotheses were supported, meeting the COnsensus-based Standards for the selection of health Measurement INstruments (COSMIN) criterion for sufficiency (≥75%) (Prinsen et al., 2018). The unsupported hypotheses concerned the correlations with the SSP and SFE-J; however, the correlation with the SFE-J was below the hypothesized range for the original PQP, suggesting that the hypothesized level itself may have been set too high. Additionally, all correlations with the original PQP exceeded 0.70. These findings suggest that the PQP-12 closely reproduces the construct-validity profile of the original PQP.

Regarding test–retest reliability, ICCs for the total score and each factor exceeded 0.70, indicating good reliability. The obtained ICCs allowed the SEM and MDC to be presented.

As for responsiveness, 80% of hypothesis tests were supported, meeting the COSMIN criterion for sufficiency (≥75%) (Prinsen et al., 2018). The only unsupported hypotheses concerned the correlation with the SDQ prosocial behavior score, which were the same unsupported hypotheses for the original PQP, indicating that the PQP-12 faithfully reproduces the responsiveness profile of the original PQP rather than losing measurement properties through shortening.

Overall, these findings suggest that the PQP-12, similar to the original PQP, can capture longitudinal changes. In COSMIN, responsiveness is defined as “the ability to detect change over time in the construct to be measured” and is an important measurement property for scales used in longitudinal and intervention research (Mokkink et al., 2018). Given that responsiveness and the MDC have not been adequately verified for several existing participation measures (Nakamura et al., 2026), it is important that the PQP-12, despite being a short form, demonstrated these indices. Because short forms of participation measures specific to children with ASD are limited, the PQP-12 is considered to enhance feasibility as a scale usable in longitudinal and intervention research while reducing respondent burden.

This study has some limitations. Although the PQP and PQP-12 are scales for children aged 3–6 years, the cohort study used to evaluate test–retest reliability and responsiveness was limited to part of the target age range. Therefore, the measurement properties of the PQP-12 require further examination in children across a broader age range. Additionally, because it is a caregiver-report scale, subjective experiences and perspectives of children may not be sufficiently reflected. These points should be investigated in future studies by applying the PQP- 12 to different age groups and combining it with other assessment methods.

## Conclusion

In this study, we developed the PQP-12, a 12-item short form of the PQP, and evaluated its measurement properties. This study revealed that the PQP-12 reduced respondent burden compared with the original PQP, multidimensionally assessed participation among children with ASD, and enhanced feasibility in large-scale surveys, longitudinal research, and clinical practice. The PQP-12 retained the four-factor structure of the original PQP and showed acceptable internal consistency, construct validity, test–retest reliability, and responsiveness. Measurement error and MDC were also presented, suggesting its usability in longitudinal and intervention research.

## Data Availability

The data that support the findings of this study are not publicly available due to ethical restrictions. Access to the data is restricted to maintain participant confidentiality as required by the ethics committee.

## Acknowledgments

We would like to express our deep gratitude to Ms. Sakumi Tachioka for her dedicated support during the preparation of this manuscript. We would also like to thank the Department of Developmental Disorders, National Institute of Mental Health, National Center of Neurology and Psychiatry (NCNP), for granting permission to use the CLASP (Check List of Obscure disAbilitieS in Preschoolers).

## Funding

This work was supported by the Japan Society for the Promotion of Science (JSPS) under KAKENHI Grant Number 22K02410 (Grant-in-Aid for Scientific Research (C)), awarded to Takuto Nakamura.

## Disclosure of Interest

The authors report no conflict of interest. While the PQP and the PQP-12 were developed with the involvement of the authors, steps to minimize bias included the use of predefined hypotheses and criteria to evaluate validity and responsiveness and adherence to standardized protocols for data collection and analysis.

## Notes

### Competing Interest Statement

The authors have declared no competing interest.

### Author Declarations

The Research Ethics Committee of Kanagawa University of Human Services gave ethical approval for this work (Approval Number: 31-14-009).

